# Finnish Diabetes Risk Score FINDRISC and HbA1c test as a screening test for the metabolic syndrome in people who lives in high-altitude, La Paz – Bolivia (3600 m.a.s.l.)

**DOI:** 10.1101/2021.03.12.21253494

**Authors:** Patricia Philco-Lima, Aydeé Cristina Ramírez-Laura, Marlene Isabel Suxo-Tejada, Ángela María Clara Alanes-Fernandez, Aida Virginia Choque-Churqui, Omar Erick Paye-Huanca, Luz Jaqueline Farah-Bravo, Carlos Ascaso-Terren

**Affiliations:** Researcher teacher’s Instituto de Investigación en Salud y Desarrollo IINSAD, Facultad de Medicina, Enfermería, Nutrición y Tecnología Médica, Universidad Mayor de San Andrés. La Paz Bolivia. Applicant in the Doctorate Program Medicina e Investigación Traslacional, Universidad de Barcelona. La Paz, Bolivia; Head of the Area of Promoción y Prevención de Enfermedades no Transmisibles y Salud Renal de la Unidad de Epidemiología del Servicio Departamental de Salud La Paz. La Paz, Bolivia; Facultad de Medicina, Enfermería, Nutrición y Tecnología Médica, Universidad Mayor de San Andrés. La Paz, Bolivia; Teacher Medicina Career, Facultad de Medicina, Enfermería, Nutrición y Tecnología Médica, Universidad Mayor de San Andrés. La Paz, Bolivia; Teacher Nutrition Career, Facultad de Medicina, Enfermería, Nutrición y Tecnología Médica, Universidad Mayor de San Andrés. La Paz, Bolivia; Biotechnology. Researcher asistant Instituto de Investigación en Salud y Desarrollo IINSAD, Facultad de Medicina, Enfermería, Nutrición y Tecnología Médica, Universidad Mayor de San Andrés. La Paz, Bolivia; Department of Basic Clinical Practice. Universitat Barcelona. Barcelona España

**Keywords:** diabetes, epidemiology of chronic non communicable diseases, health behaviour, lifestyle, health promotion

## Abstract

**BACKGROUND:** The identification of the metabolic syndrome is essential for a timely diagnosis and to avoid type 2 diabetes, the objective was to determine the validity of the FINDRISC test and glycated hemoglobin for risk screening for diabetes mellitus in a high-altitude resident population (3600 m.a.s.l.) city of La Paz - Bolivia 2018.

**METHODS:** Analytical study of diagnostic tests, the gold test was the definition of metabolic syndrome according to the criteria of the Harmonization Consensus, compared with the FINDRISC test and glycated hemoglobin. People with an age greater than or equal to 18, who agreed to participate, were included. Pregnant women, people with cancer or thyroid disease were excluded.

**RESULTS:** 400 people were included. There were 358 women (89.5%) and 42 men (10.5%) with age of 48.38±14.1 years old. The prevalence of metabolic syndrome was 46.75%. The area under the curve, sensitivity, specificity, positive predictive value and positive likelihood ratio of FINDRISC with cut-off point 10 were 0.7073(CI95% 0.66-0.75), 74.33%(CI95% 67.34-80.30), 67.14%(CI95% 60.33-73.31), 66.51%(CI95% 59.62-72.78) and 2.26(CI95% 1.83-2.79) respectively. With FINDRISC’s (cut-off point 10) and HbA1c (cut-off point 5.7%) together, were 0.8025 (CI95% 0.74-0.84), 88.1%(CI95% 80.82-92.96), 72.41%(CI95% 63.21-80.11), 77.62%(CI95% 69.74-83.98) and 3.19 (CI95% 2.36-4.32) respectively.

**CONCLUSIONS:** The FINDRISC test is a good metabolic syndrome screening test, with a cut point of 10, this is synergized with 5.7% for HbA1c in a high-altitude resident population (3600 m.a.s.l.).

**Summary Box:** What is already known on this subject?
To exist evidence that FINDRISC test is useful with HbA1c together. It to predict diabetes risk. But there are different cutoff points in both. In high altitude population as La Paz to 3600 m.a.s.l., it doesn’t know.

What are the new findings?
The FINDRISC test is a good metabolic syndrome screening test, with a cut point of 10, this is synergized with 5.7% for HbA1c in high altitude population as La Paz to 3600 m.a.s.l.

How might these results change the focus of research or clinical practice?
The FINDRISC test is easy to apply in primary care, plus the determination of glycosylated hemoglobin, enhances its diagnostic capacity to predict metabolic syndrome, in high altitude resident population, where insulin sensitivity is different.

## INTRODUCTION

Despite efforts to control the diabetes epidemic, the total number of people living with diabetes continues to rise steadily. The determination of metabolic syndrome is essential to make a timely diagnosis and take preventive measures to avoid type 2 diabetes. To detect people at risk, it is necessary to develop and implement sensitive, cost-effective and convenient detection tools to assess the diabetes risk in the primary care setting (1).

Traditionally, the Finnish Diabetes Risk Score (FINDRISC) questionnaire is considered an assessment tool to estimate the risk of type 2 diabetes, however recently it has also been used as a screening test to identify metabolic syndrome (2).

The prediction of type 2 diabetes is important for timely intervention and to avoid chronic complications associated with the disease, several studies suggest that the application of the FINDRISC test can be applied to detect metabolic syndrome in a high-risk population (3).

The simplicity and practicality of the FINDRISC test makes it more applicable to primary care, therefore it is important to measure its diagnostic capabilities against established criteria that are not as widely accepted as requiring laboratory tests.

This population of the city of La Paz has risk factors such as sedentary lifestyle, stress levels, alcohol consumption, overweight, and obesity, these characteristics make them a population at risk for type 2 diabetes.

The Finnish Diabetes Risk Score FINDRISC test was constructed from data from a Finnish population cohort of subjects aged between 35 and 64 years selected at random in 1987 and followed by 10 years, to predict the development of diabetes treated with medications. It was validated in several countries with different cut points, in South America, the use of FINDRISC is recommended, establishing the cut point at 12 as a screening method for type 2 diabetes mellitus in adults over 18 years of age in Colombia, with a strong recommendation in favor and moderate quality of evidence, data that is useful for our population (4).

The glycated hemoglobin (HbA1c) test is a blood test for type 2 diabetes and prediabetes. It measures the average level of glucose or sugar in the blood during the last three months. It can be used alone or in combination with other diabetes tests to make a diagnosis. It is also used to monitor and control diabetes.

A normal HbA1c level is less than 5.7%, prediabetes is between 5.7 to 6.4%, type 2 diabetes is above 6.5% (5).

In 2009, a harmonization conference was held to unify criteria, accepting that there was no required parameter, and waist circumference levels adapted by region and ethnicity (6). It considers the same criteria as APIII, however for the South American population a different waist circumference is taken into account, that is, greater than or equal to 90 in men and greater than or equal to 80 in women.

The study’s objective was to determine the validity of the FINDRISC test and glycated hemoglobin as a screening test to identify the risk of diabetes mellitus in the population from of city La Paz Bolivia, people who live in high-altitude (3600 m.a.s.l.).

## METHODS

It was an observational, analytical diagnostic tests study, in people of the City of La Paz, one for each urban network. The city of La Paz is 3600 meters above sea level. The data have been provided of the cross-sectional study “Prevalence of type 2 diabetes, metabolic syndrome, overweight and obesity in five merchants in the city of La Paz”, where the information of 400 people is available. The sampling was randomized probabilistic by conglomerates.

The gold standard was Metabolic Syndrome’s criteria according to the Harmonization Consensus. The comparison tests were the FINDRISC test and glycated hemoglobin.

Were included people of both sexes, with an age greater than or equal to 18 years who have accepted to participate.

We excluded women in the gestation period, people with an already defined diagnosis of some type of oncological disease, and people with a diagnosis of thyroid disease.

The FINDRISC test (4) was used as an instrument, which has 8 questions: age, body mass index, waist circumference, physical activity, diet, medication for hypertension, high glucose values, and a family history of diabetes.

For the determination of glycated hemoglobin, the immunoturbidimetric inhibition method was applied to determine the amount of hemoglobin A1c (HbA1c) as a proportion of total hemoglobin, in human whole blood (% or mmol/mol HbA1c). For this, it is necessary to release the hemoglobin contained in the red blood cells, by means of hemolysis of the sample. The absorbance of the standard and sample were read on a STAT FAX 1908+ chemical analyzer, Awareness brand, from the United States.

For the definition of the metabolic syndrome, the harmonization consensus criteria are 3 out of 5 criteria considering the waist circumference values for South Americans.

First, we compare the FINDRISC test with a cut-off point of 10, 12, 13, and 15 against the definition of metabolic syndrome according to the harmonization consensus as the gold standard, then using a ROC curve to find the best FINDRISC cutoff point. The same analysis was performed with HbA1c with 5.7%, 5.9%, and 6.5% and with the better cut-off FINDRISC test and the better cut-off point HbA1c together.

To compare the roc curves, it was possible to do this using the logistic linear predictors and the roccomp command in Stata.

Then 2×2 tables were constructed for the calculation of sensitivity, specificity, positive and negative predictive values, and positive and negative likelihood ratios.

The study from which this research derives has the ethical endorsement of the Research Ethics Committee – IINSAD. This certificate has as date November 17th, 2017.

## RESULTS

Information was obtained from 400 people, who met the measurements of the metabolic syndrome criteria. There were 358 women (89.5%) and 42 men (10.5%), with an average age of 48.38 ± 14.1 years, with a minimum age of 18 years and a maximum of 84.6 years.

The prevalence of metabolic syndrome was 46.75% IC_95%_ (41.77-51.77).

The Finnish Diabetes Risk Score FINDRISC diagnostic features obtained with the cut-off points 10, 12, 13, and 15 there were in the table N°1, where the sensivility was better with cut-off point 10.

In relation to glycated hemoglobin, its median was 5.85; RIC: 5.4-6.3%, minimum of 4.5%, and maximum 11.8%. The diagnostic features obtained with the cut-off point of 5.7% in comparison to 5.9% are the same (Table 2).

**Table 1.**
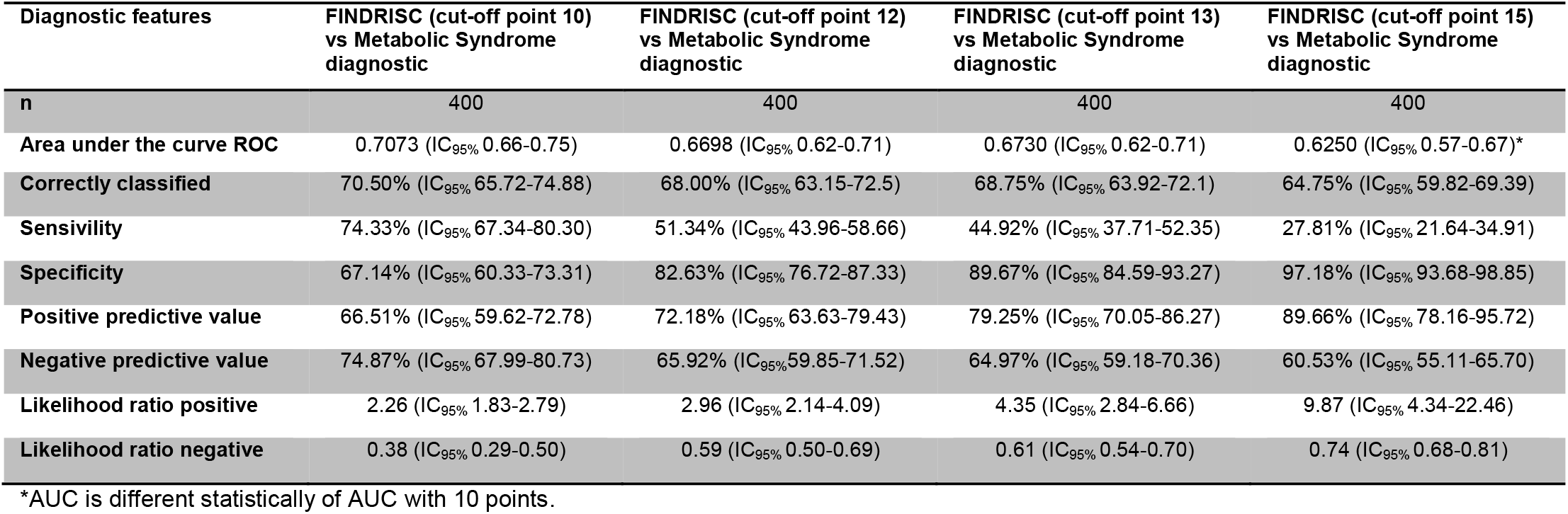
Differences between FINDRISC with diagnostic features of 10 points and cut-off point recommended (12, 13 y 15 points)

| Diagnostic features | FINDRISC (cut-off point 10)<br>vs Metabolic Syndrome<br>diagnostic | FINDRISC (cut-off point 12)<br>vs Metabolic Syndrome<br>diagnostic | FINDRISC (cut-off point 13)<br>vs Metabolic Syndrome<br>diagnostic | FINDRISC (cut-off point 15)<br>vs Metabolic Syndrome<br>diagnostic |
| --- | --- | --- | --- | --- |
| n | 400 | 400 | 400 | 400 |
| Area under the curve ROC | 0.7073 (IC<sub>95%</sub> 0.66-0.75) | 0.6698 (IC<sub>95%</sub> 0.62-0.71) | 0.6730 (IC<sub>95%</sub> 0.62-0.71) | 0.6250 (IC<sub>95%</sub> 0.57-0.67)* |
| Correctly classified | 70.50% (IC<sub>95%</sub> 65.72-74.88) | 68.00% (IC<sub>95%</sub> 63.15-72.5) | 68.75% (IC<sub>95%</sub> 63.92-72.1) | 64.75% (IC<sub>95%</sub> 59.82-69.39) |
| Sensivity | 74.33% (IC<sub>95%</sub> 67.34-80.30) | 51.34% (IC<sub>95%</sub> 43.96-58.66) | 44.92% (IC<sub>95%</sub> 37.71-52.35) | 27.81% (IC<sub>95%</sub> 21.64-34.91) |
| Specificity | 67.14% (IC<sub>95%</sub> 60.33-73.31) | 82.63% (IC<sub>95%</sub> 76.72-87.33) | 89.67% (IC<sub>95%</sub> 84.59-93.27) | 97.18% (IC<sub>95%</sub> 93.68-98.85) |
| Positive predictive value | 66.51% (IC<sub>95%</sub> 59.62-72.78) | 72.18% (IC<sub>95%</sub> 63.63-79.43) | 79.25% (IC<sub>95%</sub> 70.05-86.27) | 89.66% (IC<sub>95%</sub> 78.16-95.72) |
| Negative predictive value | 74.87% (IC<sub>95%</sub> 67.99-80.73) | 65.92% (IC<sub>95%</sub> 59.85-71.52) | 64.97% (IC<sub>95%</sub> 59.18-70.36) | 60.53% (IC<sub>95%</sub> 55.11-65.70) |
| Likelihood ratio positive | 2.26 (IC<sub>95%</sub> 1.83-2.79) | 2.96 (IC<sub>95%</sub> 2.14-4.09) | 4.35 (IC<sub>95%</sub> 2.84-6.66) | 9.87 (IC<sub>95%</sub> 4.34-22.46) |
| Likelihood ratio negative | 0.38 (IC<sub>95%</sub> 0.29-0.50) | 0.59 (IC<sub>95%</sub> 0.50-0.69) | 0.61 (IC<sub>95%</sub> 0.54-0.70) | 0.74 (IC<sub>95%</sub> 0.68-0.81) |

\*AUC is different statistically of AUC with 10 points.

**Table 2.**
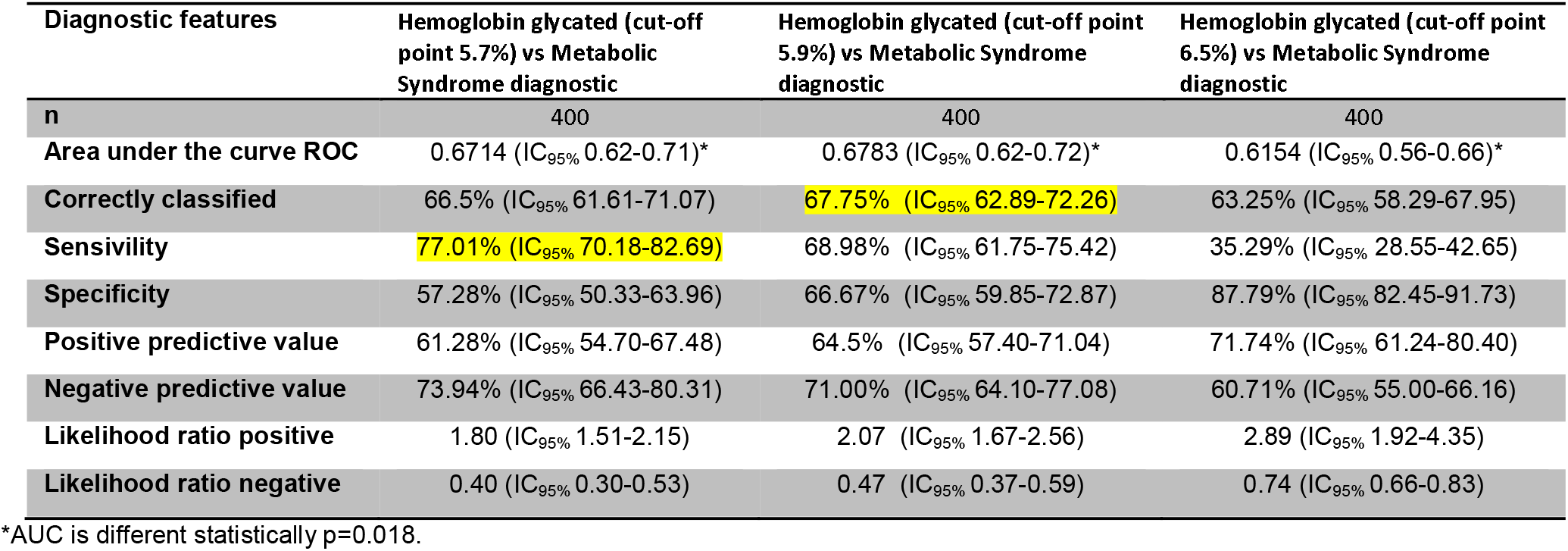
Differences between hemoglobin glycated’s diagnostic features with 5.7%, 5.9% and 6.5%.

| Diagnostic features | Hemoglobin glycated (cut-off point 5.7%) vs Metabolic Syndrome diagnostic | Hemoglobin glycated (cut-off point 5.9%) vs Metabolic Syndrome diagnostic | Hemoglobin glycated (cut-off point 6.5%) vs Metabolic Syndrome diagnostic |
| --- | --- | --- | --- |
| n | 400 | 400 | 400 |
| Area under the curve ROC | 0.6714 (IC<sub>95%</sub> 0.62-0.71)* | 0.6783 (IC<sub>95%</sub> 0.62-0.72)* | 0.6154 (IC<sub>95%</sub> 0.56-0.66)* |
| Correctly classified | 66.5% (IC<sub>95%</sub> 61.61-71.07) | 67.75% (IC<sub>95%</sub> 62.89-72.26) | 63.25% (IC<sub>95%</sub> 58.29-67.95) |
| Sensitivity | 77.01% (IC<sub>95%</sub> 70.18-82.69) | 68.98% (IC<sub>95%</sub> 61.75-75.42) | 35.29% (IC<sub>95%</sub> 28.55-42.65) |
| Specificity | 57.28% (IC<sub>95%</sub> 50.33-63.96) | 66.67% (IC<sub>95%</sub> 59.85-72.87) | 87.79% (IC<sub>95%</sub> 82.45-91.73) |
| Positive predictive value | 61.28% (IC<sub>95%</sub> 54.70-67.48) | 64.5% (IC<sub>95%</sub> 57.40-71.04) | 71.74% (IC<sub>95%</sub> 61.24-80.40) |
| Negative predictive value | 73.94% (IC<sub>95%</sub> 66.43-80.31) | 71.00% (IC<sub>95%</sub> 64.10-77.08) | 60.71% (IC<sub>95%</sub> 55.00-66.16) |
| Likelihood ratio positive | 1.80 (IC<sub>95%</sub> 1.51-2.15) | 2.07 (IC<sub>95%</sub> 1.67-2.56) | 2.89 (IC<sub>95%</sub> 1.92-4.35) |
| Likelihood ratio negative | 0.40 (IC<sub>95%</sub> 0.30-0.53) | 0.47 (IC<sub>95%</sub> 0.37-0.59) | 0.74 (IC<sub>95%</sub> 0.66-0.83) |

\*AUC is different statistically p=0.018.

Combined the better cut-off point of FINDRISC and Hba1c together, we compared 4 groups FINDRISC with 10 points, and HbA1c with 5.7%, FINDRISC with 10 points, and HbA1c with 5.9%, FINDRISC with 12 points, and HbA1c with 5.7%, and FINDRISC with 12 points, and HbA1c with 5.9% (Table 3). The AUC is the same, however, FINDRISC with 10 points, and HbA1c with 5.7% had better sensivility.

**Table 3.** Diagnostic features between FINDRISC plus hemoglobin glycated vs metabolic syndrome diagnostic.

| Diagnostic features | FINDRISC (cut-off point 10) and hemoglobin glycated (cut-off point 5.7%) together vs Metabolic Syndrome diagnostic | FINDRISC (cut-off point 10) and hemoglobin glycated (cut-off point 5.9%) together vs Metabolic Syndrome diagnostic | FINDRISC (cut-off point 12) and hemoglobin glycated (cut-off point 5.7%) together vs Metabolic Syndrome diagnostic | FINDRISC (cut-off point 12) and hemoglobin glycated (cut-off point 5.9%) together vs Metabolic Syndrome diagnostic |
| --- | --- | --- | --- | --- |
| n | 242 | 241 | 216 | 229 |
| Area under the curve ROC | 0.8025 (IC<sub>95%</sub> 0.74-0.84) | 0.8189 (IC<sub>95%</sub> 0.76-0.86) | 0.8160 (IC<sub>95%</sub> 0.75-0.86) | 0.7985 (IC<sub>95%</sub> 0.74-0.84) |
| Prevalence of metabolic syndrome | 52.07% (IC<sub>95%</sub> 45.58-58.48) | 47.72% (IC<sub>95%</sub> 41.29-54.22) | 46.76% (IC<sub>95%</sub> 39.99-53.64) | 43.67% (IC<sub>95%</sub> 37.19-50.36) |
| Correctly classified | 80.58% (IC<sub>95%</sub> 74.91-85.25) | 81.74% (IC<sub>95%</sub> 76.16-86.29) | 81.94% (IC<sub>95%</sub> 76.02-86.70) | 81.22% (IC<sub>95%</sub> 75.43-85.94) |
| Sensitivity | 88.1% (IC<sub>95%</sub> 80.82-92.96) | 85.22% (IC<sub>95%</sub> 77.10-90.91) | 76.24% (IC<sub>95%</sub> 66.54-83.90) | 69.00% (IC<sub>95%</sub> 58.85-77.66) |
| Specificity | 72.41% (IC<sub>95%</sub> 63.21-80.11) | 78.57% (IC<sub>95%</sub> 70.19-85.17) | 86.96% (IC<sub>95%</sub> 79.09-92.27) | 90.70% (IC<sub>95%</sub> 83.98-94.89) |
| Positive predictive value | 77.62% (IC<sub>95%</sub> 69.74-83.98) | 78.40% (IC<sub>95%</sub> 69.96-85.05) | 83.70% (IC<sub>95%</sub> 74.21-90.29) | 85.19% (IC<sub>95%</sub> 75.16-91.78) |
| Negative predictive value | 84.85% (IC<sub>95%</sub> 75.92-90.99) | 85.34% (IC<sub>95%</sub> 77.29-90.99) | 80.65% (IC<sub>95%</sub> 72.37-86.97) | 79.05% (IC<sub>95%</sub> 71.44-85.12) |
| Likelihood ratio positive | 3.19 (IC<sub>95%</sub> 2.36-4.32) | 3.98 (IC<sub>95%</sub> 2.82-5.60) | 5.84 (IC<sub>95%</sub> 3.6-9.49) | 7.42 (IC<sub>95%</sub> 4.26-12.92) |
| Likelihood ratio negative | 0.16 (IC<sub>95%</sub> 0.1-0.27) | 0.19 (IC<sub>95%</sub> 0.12-0.29) | 0.27 (IC<sub>95%</sub> 0.19-0.39) | 0.34 (IC<sub>95%</sub> 0.25-0.46) |

## DISCUSSION

The prevalence of metabolic syndrome is high (46.7%), higher than that finded in CARMELA study, which was 42%, the highest in South America (7). In other words, the population sample has almost 50% of people who are at risk of having diabetes, cardiovascular disease, or cancer in the future. Screening strategies are implemented in several countries to identify subjects at risk of diabetes mellitus.

Several statistical models have been generated to predict type 2 diabetes or cardiovascular disease, instruments constructed from risk factors such as age, sex, obesity, diet, physical activity, ethnicity, family history of diabetes, etc., one of these instruments is FINDRISC, however, with the recommended score in Finland, does not have predictive precision in Latin Americans, the biochemical models being superior (8), which is why considering the FINDRISC risk score plus glycated hemoglobin is the best option to make the prediction in Latinos. However, the physical condition and physiological effects derived from living at high altitudes (3600 m.a.s.l.), are necessary to determine if this cut off point is different.

The FINDRISC test has been used to identify the risk of type 2 diabetes since 1987, it is probably one of the most efficient to identify this risk, it is considered a rapid test, with easy applicability. It was validated in several countries with different cut-off points, in Europe with 15 and in South America with 12.

High-altitude hypoxia improves insulin sensitivity which is associated with adenosine monophosphate-activated protein kinase AMPK activation in the skeletal muscle and consequently enhanced mitochondrial biogenesis and fatty acid oxidation (9). For this reason, is important to find the FINDRISC and HbA1c cut off correct for population resident to high altitude.

Internationally HbA1c values between 5.7 and 6.4 % identify prediabetic individuals by Asociación Latinoamericana de Diabetes ALAD (10). But our results differ, it is recommended to use the cut-off point ≥ 10 points FINDRISC test and the cut-off point ≥5.7% glycated hemoglobin in population resident in high altitude.

In relation FINDRISC test, there are no statistically significant differences between the curves with the cut-off point 10, 12 and 13 points to predict metabolic syndrome, it is considered that the best point is with 10 points for having the highest sensitivity and highest number of correct answers (70.5%), better than 12 points, which is the one proposed for the South American population (4).

The differences between the areas with 10 and 15 points is significant statistically (p=0.0006) (Table 1) which corresponds to the smallest area under the curve, that is, 15 is the worst result.

FINDRISC test with cut-off point of 10 to have almost 3.9 times more the probability to be positive, when the test is positive and this effect differs than the situation with 12, 13 or 15.

In Belgians (3) with cut-off 13, they found an area under the curve ROC 0.76 (IC95% 0.72-0.82), the sensitivity of 64%, and specificity 63%, all lower values than our values.

In Iranian people, with cut-off 12, the area under the curve ROC was del 65.0% (95% CI 61.3-68.7) (11), lower values in relation to this study.

In relation to glycosylated hemoglobin, to predict diabetes or metabolic syndrome, glycosylated hemoglobin has been shown to be better than fasting blood glucose, there is evidence that the value of glycated hemoglobin increases on average 0.2% in patients with metabolic syndrome (9). In the present study, AUC is the same with 5.7% and 5.9% HbA1c, but both are different of 6.5% cut off point. Since there are no significant differences between the AUC with 5.7% and 5.9% cutoff points, it is recommended to keep the 5.7% point as its sensivility is the best 77.01% (IC95% 70.18-82.69) though it is low in front of 82.6% sensivility with 5.9% in the study of E. Martin, E. Ruf, R. Landgraf, et al (1) and specificity 64.9%, they found with ROC analyses that the optimal cut off levels was for FINDRISC ≥ 12 points and for HbA1c ≥ 5.9 %.

The diagnostic capacity of the FINDRISC test is synergized with glycated hemoglobin in people who live at high altitude. A similar result was obtained by Meijnikman (3) he found that with 13 as cutoff point the AUC was 0.76 (95% CI 0.72–0.82), sensitivity was 64% and specificity was 63% adding a determination of glycated hemoglobin, increased the area under the curve significantly to 0.93 (CI95% 0.9-0.97). In high altitude adding HbA1c, the curve increase to 0.80 (IC95% 0.74-0.84) whit a sensitivity of 88.1% and specificity was 72.41%.

In conclusion, diagnostic features of the FINDRISC test in relation to the definition of metabolic syndrome according to the Harmonization Consensus, indicate it as a good screening test with a cut-off point of 10 for people that live in high altitude conditions. The FINDRISC test with a cut-off point of 10 and hemoglobin glycated with a cut-off point of 5.7% together, they do a synergize effect as a screening test in the population who lives in high altitude.

Accessible diagnostics enhances the quality of care in areas with limited laboratory infrastructure. It is useful to ensure healthy lives, promote well-being for all, and catch the universal healthcare 2030, it helps to Reduce inequality within and among countries, as recommended in The-Sustainable-Development-Goals (12).

Among the limitations, it can be mentioned that the population comes from the commercial sector and that a large part of the sample was women, so it is recommended that more research be carried out to confirm these results.

## Data Availability

It is declared that all the data referred to in the manuscript and the links below are available.

